# Impact of untreated periodontal disease, periodontal treatment, and regular dental prophylaxis on chronic kidney disease outcomes

**DOI:** 10.64898/2026.05.09.26352792

**Authors:** Scott G. Silvey, Janina Golob Deeb, Jasmohan S. Bajaj, Nilang Patel

## Abstract

**Rationale & Objective:** Periodontal disease(PD), a chronic inflammatory condition, may contribute to chronic kidney disease(CKD) through systemic inflammation, but its impact on CKD outcomes and the potential protective effects of periodontal treatment and routine dental prophylaxis remain uncertain. This study evaluated associations between PD, dental interventions, and kidney outcomes in a large U.S. veteran cohort.

**Study Design:** Retrospective cohort study.

**Setting & Participants:** Using Veterans Health Administration data(2009–2019), we identified 86,376 adults eligible for comprehensive dental care with baseline eGFR >60 mL/min/1.73m² and followed them from their initial dental examination.

**Exposure:** Patients with PD (Cohort-A) were divided into those who received periodontal treatment (PD-Treated), those who did not receive treatment but had ≥1 dental prophylaxis visit/year (PD-Prophylaxis), and those who received neither (PD-Untreated). Those without PD (Cohort-B) were grouped by presence or absence of regular dental prophylaxis (≥1 visit/year).

**Outcome:** Incident CKD (eGFR <60 mL/min/1.73 m² and >25% decline from baseline), ≥40% eGFR reduction, and incident albuminuria (>30 mg/g), each confirmed with repeat labs ≥90 days apart.

**Analytical Approach:** Multivariable logistic regression model

**Results:** Of 86,376 veterans (mean age 57.17±12.59 years; 91.4% male), 37.6% had PD. Adjusted model showed significantly lower odds of incident CKD, ≥40% eGFR decline, and incident albuminuria noted in both PD-Treated [OR(95%CI): 0.80(0.70-0.91), 0.69(0.56-0.84), 0.88 (0.79-0.99)] and PD-Prophylaxis groups [OR(95%CI): 0.81(0.66-0.98), 0.60(0.43-0.82), 0.79(0.67-0.94)] compared to the PD-Untreated. Similarly, among patients without PD, regular dental prophylaxis was associated with reduced odds of Incident CKD, ≥40% eGFR decline, and incident albuminuria [OR(95%CI): 0.87(0.78-0.96), 0.76(0.65-0.90), 0.85(0.78-0.93)].

**Limitations:** Retrospective design, unmeasured confounders, and reliance on electronic health records.

**Conclusions:** PD is associated with increased risks of incident CKD, accelerated eGFR decline, and new-onset albuminuria. Periodontal treatment and routine dental prophylaxis mitigate these risks. Even in individuals without PD, regular dental prophylaxis appears protective against CKD development and progression.

## Introduction

Periodontal disease (PD) represents a significant public health concern, affecting approximately 50% of US adults, with a prevalence increasing with age and thus higher among elderly, and those who do not receive prophylactic dental care.[1,2] This chronic inflammatory condition affects the supporting structures of teeth and is gaining attention as a potential systemic health risk beyond the oral cavity.

Emerging evidence suggests a bidirectional relationship between PD and various systemic conditions, including diabetes mellitus, cardiovascular disease, and chronic kidney disease (CKD).[3–8]

The mechanistic underpinnings of this relationship are multifaceted, involving the systemic dissemination of oral pathogens and their byproducts, which subsequently induce low-grade systemic inflammation and endothelial dysfunction.[9] This pathophysiological cascade suggests that modulation of PD could potentially serve as a supportive therapy in mitigating CKD progression risk.

Regular dental prophylaxis provided at the dental office encompassing the removal of dental plaque (biofilm) and calculus, identifying and modifying local factors facilitating plaque accumulation, identifying and managing systemic risk factors, and assessing and improving patients’ oral hygiene habits and efficiency at home, is fundamental in establishing optimal oral health and preventing and maintaining PD. Extrapolating from this, it can be postulated that effective management of PD, coupled with consistent and regular dental prophylaxis and periodontal maintenance, may attenuate the systemic inflammatory burden and, consequently, the risk of CKD progression. However, there is a lack of strong empirical evidence supporting this hypothesis.

The objective of the current study is to examine the relationship between periodontal disease (PD), its treatment, presence, absence and frequency of dental prophylaxis, and the development and progression of CKD in a cohort of veterans with normal kidney function who are eligible for continuous and comprehensive dental coverage provided by the VA. We hypothesized that treatment of periodontal disease and its prevention through regular dental prophylaxis may reduce the incidence and progression of chronic kidney disease.

## Methods

Using the VA Corporate Data Warehouse (VHA-CDW), veterans eligible for continuous and comprehensive dental care for 5 consecutive years at the VA from 2009-2014 were identified and followed up from the date of their initial (index) comprehensive dental exam (CDT code: D0150) until December 31, 2019. Veterans who had a dental exam in the previous 3 years or were edentulous during the initial comprehensive exam were excluded. Patients with baseline eGFR > 60 mL/min/1.73m^2^, defined as averaging values ± 6 months from index comprehensive exam and no prior evidence of dialysis/transplant were followed. eGFR was calculated using the 2021 CKD-EPI race free equation.[10]

Baseline demographics, social-economic data, comorbidities, medications, labs and dental treatment records were obtained. We selected these variables based on common dental health risk factors such as alcohol and tobacco use, as well as social determinants of health like median household income, urban vs. rural residence, homelessness, and educational status as well as kidney disease risk factors such as Diabetes, Hypertension and NSAIDs and PPI usage, to account for potential cofounders that could modify the effect of dental prophylaxis on outcomes. (Details in Supplement S1 Table).

Patients were divided into two cohorts; Cohort A: Those diagnosed with PD within 1 year of index exam date and Cohort B: those without PD. PD was diagnosed using validated ICD-9/10 codes (Supplement S1 Table). Cohort A was divided into three groups based on the presence or absence of periodontal treatment (as indicated by ADA CDT codes: D4341, D4342, D4910) and regular dental prophylaxis (defined as ≥ 1 dental prophylaxis visit per year based on CDT code: D1110 or D4355). Those who received periodontal treatment (PD-Treated), those who did not receive periodontal treatment but received regular dental prophylaxis (PD-Prophylaxis), and those who received neither (PD-Untreated).

Patients in Cohort B were divided into two groups based on presence or absence of regular dental prophylaxis as defined earlier (> 1 dental prophylaxis visit per year or < 1 dental prophylaxis visit per year) (Fig 1)

**Fig 1:**
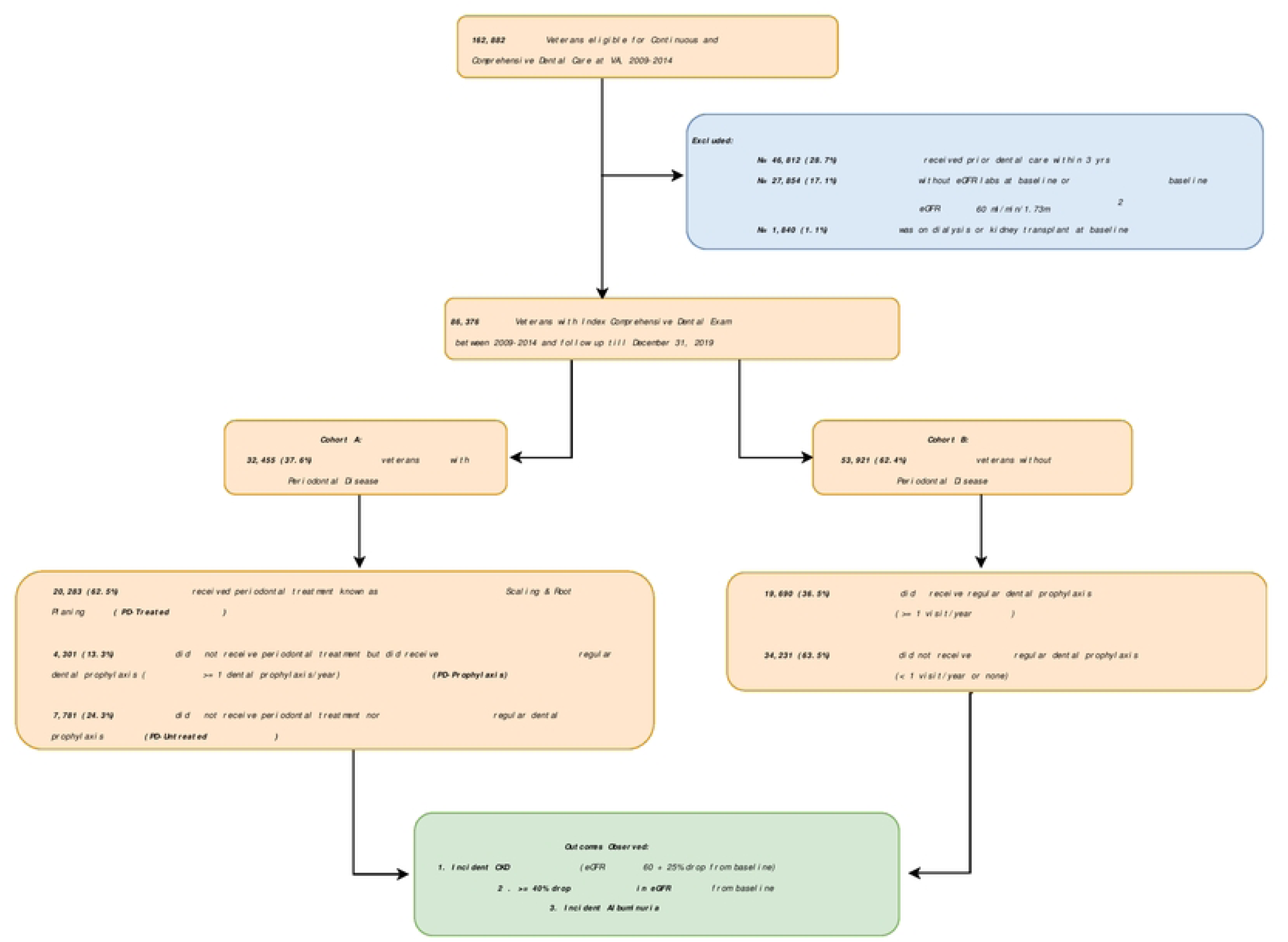
Flowchart

### Analysis of eGFR Labs

Follow-up eGFR labs were analyzed starting one year after the initial exam until the end of the observation period (December 31, 2019). The primary outcome was the development of CKD (Incident CKD), defined as an eGFR below 60 mL/min/1.73m² that was more than 25% lower than baseline. A second confirmatory lab was required at least 90 days after the first instance. As a secondary outcome, we considered a ≥ 40% reduction in eGFR from baseline, also confirmed ≥ 2 times 90 days apart.

### Analysis of UACR Labs

Within these patients, we also analyzed urine albumin/creatinine ratio (UACR) labs when available. Baseline UACR was calculated by averaging all labs within ±1 year of the index exam. Those with baseline UACR > 30 mg/gm, indicative of albuminuria at baseline, were removed. For those without baseline UACR value, urine dipstick or UPCR results were converted based on published literature.[11] We analyzed the odds of new-onset albuminuria (Incident Albuminuria), defined as UACR > 30 mg/gm during follow-up, using logistic regression analysis, requiring a second confirmatory lab ≥ 90 days later.

### Statistical Analysis

Characteristics between the groups within Cohort A/B were compared using one-way Anova, two sample *t*-tests, or Chi-Squared tests as appropriate; variables collected included demographics, medications, comorbidities, and social-economic variables. Logistic regression models were used to compare the odds of these outcomes between the groups within cohorts A/B. For all models, we also adjusted for 1.) all covariates that were significantly different between the groups and 2.) follow-up time in days, since each patient was followed for a different number of days post-index exam. Final logistic regression models were evaluated for multicollinearity using variance inflation factors (VIF), with values > 5 indicating notable correlation between predictors.[12]

As a sensitivity analysis, we also included baseline UACR (if available) as a covariate to be adjusted for in multivariable models.

RStudio version 4.5.2. (Posit) was used for all statistical analyses. All hypothesis tests conducted were 2-sided, with statistical significance defined as *P* < .05.

This cohort study was approved by the institutional review board of the Central Virginia VA Healthcare System (ID:02600). Data including identifiable information were accessed for research purpose between February 2025 to October 2025 on the Department of Veterans Affairs (VA) Informatics and Computing Infrastructure (VINCI) . All the analysis were done on the secure VINCI computing platform which stays behind VA Firewall. Only non-identifiable aggregate result were downloaded for study reporting. A waiver of informed consent was granted because the study was a retrospective analysis. The study followed the Strengthening the Reporting of Observational Studies in Epidemiology (<u>STROBE</u>) reporting guideline.

## Results

Final cohort contained 86,376 patients (avg age 57.17 ± 12.59 years, 91.4% male). Of these patients, 32,455(37.6%) had PD(Cohort A), and 53,921(62.4%) did not have PD(Cohort B) (Fig 1). Patients with PD were generally older, more likely male, and had a higher BMI. They were more often of Hispanic or Latino ethnicity, races other than white, lived in urban areas, and had a greater burden of comorbidities, as well as higher rates of alcohol and tobacco use disorders. (S2 Table) The median number of labs used to calculate baseline eGFR was 13 (IQR: 8-21).

### Cohort A

Among individuals with periodontal disease (avg age 59.02±10.61 years, 94.1% male), 62.5% underwent periodontal treatment(PD-Treated). Of the remaining, 4301(13.3%) were not receiving periodontal treatment but received regular dental prophylaxis(PD-Prophylaxis) while 7871(24.3%) had neither periodontal treatment nor yearly dental prophylaxis(PD-Untreated). (Fig 1)

Patients receiving periodontal treatment tended to be male, often of Hispanic ethnicity, and likely to live in urban areas. In contrast, those who did not receive periodontal treatment nor regular dental prophylaxis(PD-Untreated) tended to reside in lower-income neighborhoods with fewer bachelor’s degrees, had a higher smoking history, greater comorbidity burden, increased cardiovascular disease risk, and higher statin use.(Table 1)

**Table 1.** Cohort A, Periodontal Disease (*n* = 32,455)

|  | **Periodontal Disease with Periodontal Treatment (PD-Treated) (n = 20283, 62.5%)** | **Periodontal Disease without Periodontal Treatment, with Regular Dental Prophylaxis (>=1 visit/year) (PD-Prophylaxis) (n = 4301, 13.3%)** | **Periodontal Disease with neither Periodontal Treatment nor Regular Dental Prophylaxis (< 1 visit/year) (PD-Untreated) (n = 7871, 24.3%)** | **p-value** |
| --- | --- | --- | --- | --- |
| **Variable, Mean ± SD or n (%)** |  |  |  |  |
| **Demographics** |  |  |  |  |
| Age (> 65) | 5042 (24.9%) | 1255 (29.2%) | 1944 (24.7%) | **<0.001** |
| Male Gender | 19178 (94.6%) | 3995 (92.9%) | 7368 (93.6%) | **<0.001** |
| White Race | 13731 (74.3%) | 3189 (80.3%) | 5418 (74.5%) | **<0.001** |
| BMI (> 30) | 10967 (54.1%) | 2296 (53.4%) | 3967 (50.4%) | **<0.001** |
| Hispanic or Latino Ethnicity | 1841 (9.4%) | 289 (6.9%) | 581 (7.6%) | **<0.001** |
| **Social Determinants of Health** |  |  |  |  |
| Median Household Income | \$52423 (\$17991) | \$52836 (\$18760) | \$50803 (\$17675) | **<0.001** |
| Urban Residence | 9589 (62.3%) | 1921 (60.2%) | 3645 (59.8%) | **<0.001** |
| Homelessness | 214 (1.4%) | 38 (1.2%) | 105 (1.7%) | 0.085 |
| % with Bachelor's Degree | 23.78 (12.23) | 24.48 (12.54) | 22.97 (12.09) | **<0.001** |
| Current or Former Smoker | 10663 (69.9%) | 1969 (64.1%) | 4599 (74.3%) | **<0.001** |
| Alcohol Use Disorder | 3662 (18.1%) | 577 (13.4%) | 1358 (17.3%) | **<0.001** |
| **Comorbidities and Lab** |  |  |  |  |
| Diabetes | 6835 (33.7%) | 1384 (32.2%) | 2634 (33.5%) | 0.159 |
| Hypertension | 13208 (65.1%) | 2815 (65.4%) | 5191 (66.0%) | 0.417 |
| CVD including Heart Failure | 3760 (18.5%) | 925 (21.5%) | 1753 (22.3%) | **<0.001** |
| Charlson Comorbidity Index (> 3) | 1447 (7.1%) | 340 (7.9%) | 660 (8.4%) | **0.001** |
| Baseline eGFR (ml/min/1.73m<sup>2</sup>) | 87.85 (14.18) | 86.97 (13.57) | 88.02 (14.25) | **<0.001** |
| **Medications** |  |  |  |  |
| ACE-I/ARB | 7490 (36.9%) | 1634 (38.0%) | 2955 (37.5%) | 0.335 |
| Statin | 4534 (22.4%) | 967 (22.5%) | 1836 (23.3%) | 0.211 |
| PPI | 2872 (14.2%) | 674 (15.7%) | 1120 (14.2%) | **0.034** |
| NSAIDs | 3057 (15.1%) | 676 (15.7%) | 1207 (15.3%) | 0.535 |
| **Outcomes** |  |  |  |  |
| Incident CKD | 1470 (7.2%) | 351 (8.2%) | 712 (9.0%) | **<0.001** |
| ≥40% eGFR decline | 529 (2.6%) | 114 (2.7%) | 286 (3.6%) | **<0.001** |
| Incident Albuminuria* | 2434 (18.4%) | 482 (16.5%) | 979 (19.8%) | **0.001** |

\*Among those who had valid UACR labs available.
BMI- Body Mass Index, ACE-I/ARB – Angiotensin Converting Enzyme Inhibitors or Angiotensin Receptor Blockers, PPI- Proton Pump Inhibitors, NSAIDS - Nonsteroidal Anti-Inflammatory Drugs, CCI - Charlson Comorbidity Index, CVD – Cardiovascular Disease, CKD-Chronic Kidney Disease

### Cohort B

Among individuals without periodontal disease (avg age 56.06±13.52 years, 89.8% male), 36.5% received regular dental prophylaxis(≥ 1 visits per year). Regular dental prophylaxis patients were older, more likely to be White or Hispanic, and had a higher body mass index (BMI). They lived in neighborhoods with higher median household incomes and more residents holding at least a bachelor’s degree. They had lower charlson comorbidity index(CCI), smoking history, and alcohol use disorder but were more likely to have hypertension or be on ACE-I/ARB.(Table 2)

**Table 2.** Cohort B, No Periodontal Disease (*n* = 53921)

|  | **Regular Dental Prophylaxis**<br>**(≥ 1 visit/year)**<br>**(n =19690, 36.5%)** | **No Regular Dental Prophylaxis**<br>**(< 1 visit/year)**<br>**(n = 34231, 63.5%)** | **p-value** |
| --- | --- | --- | --- |
| **Variable, Mean ± SD or n (%)** |  |  |  |
| **Demographics** |  |  |  |
| Age (> 65) | 5245 (26.6%) | 7897 (23.1%) | **<0.001** |
| Male Gender | 17675 (89.8%) | 30760 (89.9%) | 0.740 |
| White Race | 14613 (80.7%) | 24510 (78.0%) | **<0.001** |
| BMI (> 30) | 10593 (53.8%) | 17144 (50.1%) | **<0.001** |
| Hispanic or Latino Ethnicity | 1510 (7.9%) | 2421 (7.3%) | **0.013** |
| **Social Determinants of Health** |  |  |  |
| Median Household Income | \$53040 (\$18394) | \$51228 (\$17649) | **<0.001** |
| Urban Residence | 8564 (58.4%) | 15126 (57.6%) | 0.132 |
| Homelessness | 240 (1.6%) | 383 (1.5%) | 0.173 |
| % with Bachelor's Degree | 24.66 (12.48) | 23.30 (12.22) | **<0.001** |
| Current or Former Smoker | 7935 (57.2%) | 18247 (68.5%) | **<0.001** |
| Alcohol Use Disorder | 2513 (12.8%) | 5567 (16.3%) | **<0.001** |
| **Comorbidities and Lab** |  |  |  |
| Diabetes | 5672 (28.8%) | 9995 (29.2%) | 0.339 |
| Hypertension | 11942 (60.7%) | 19800 (57.8%) | **<0.001** |
| CVD including Heart Failure | 3635 (18.5%) | 6289 (18.4%) | 0.807 |
| Charlson Comorbidity Index (> 3) | 1229 (6.2%) | 2575 (7.5%) | **<0.001** |
| Baseline eGFR (ml/min/1.73m2) | 88.36 (14.49) | 89.96 (15.53) | **<0.001** |
| **Medications** |  |  |  |
| ACE-I/ARB | 6794 (34.5%) | 10919 (31.9%) | **<0.001** |
| Statin | 4294 (21.8%) | 6597 (20.3%) | **<0.001** |
| PPI | 3062 (15.6%) | 5168 (15.1%) | 0.162 |
| NSAIDs | 3014 (15.3%) | 5338 (15.6%) | 0.382 |
| **Outcomes** |  |  |  |
| Incident CKD | 1297 (6.6%) | 2596 (7.6%) | **<0.001** |
| ≥40% eGFR decline | 400 (2.0%) | 979 (2.9%) | **<0.001** |
| Incident Albuminuria* | 1963 (15.0%) | 3814 (17.4%) | **<0.001** |

\*Among those who had valid UACR labs available.
BMI- Body Mass Index, ACE-I/ARB – Angiotensin Converting Enzyme Inhibitors or Angiotensin Receptor Blockers, PPI- Proton Pump Inhibitors, NSAIDS - Nonsteroidal Anti-Inflammatory Drugs, CCI - Charlson Comorbidity Index, CVD – Cardiovascular Disease, CKD-Chronic Kidney Disease

### Analysis of eGFR Labs

Among 86,376 patients, a total of 1,556,798 labs were collected over the observation period. The median number of labs per-patient was 14 (IQR: 9-22) with an average follow-up time of 7 years (IQR: 6-9 years). During this period, 6426 (7.4%) developed new-onset CKD and 2308 (2.7%) had an absolute eGFR drop >40% of baseline.

### Cohort A

#### Incident CKD

People with periodontal disease who received treatment(PD-Treated) or had at least one dental prophylaxis visit per year(PD-Prophylaxis) had significantly lower CKD incidence compared to those without treatment and less frequent dental prophylaxis visits(PD-Untreated). (Table 1).

Those receiving periodontal treatment(PD-Treated) had 21% lower CKD risk (95% CI: [0.72-0.86], p<0.001), and those receiving regular dental prophylaxis(PD-Prophylaxis) had 11% lower CKD risk (95% CI: [0.78-1.02], p=0.098), compared to those of PD-Untreated group. This trend was not statistically significant in univariable analysis (Table 3). After adjusting for confounding variables, the association remained consistent for PD-Treated group (PD-Treated vs. PD-Untreated; OR: 0.80 [0.70-0.91], p<0.001), and PD-Prophylaxis vs. PD-Untreated became significant (OR: 0.81 [0.66-0.98], p=0.030) (Fig 2). Other variables associated with new-onset CKD were older age, non-white race, higher BMI, current or former smoking, higher CCI, Cardiovascular Disease (CVD) including Heart Failure, statin, and proton pump inhibitors(PPI) usage (S3 Table).

**Table 3:** Univariable and Multivariable analysis of outcomes.

| Cohort A – Patients with Periodontal Disease |  |  |  |  |  |
| --- | --- | --- | --- | --- | --- |
|  | Univariable Analysis |  |  | Multivariable Analysis* |  |
| Outcome | Odds Ratio (95% CI) | p-value |  | Odds Ratio (95% CI) | p-value |
| **Incident CKD** |  |  |  |  |  |
| PD-Treated vs. PD-Untreated | 0.79 [0.72-0.86] | **<0.001** |  | 0.80 [0.70-0.91] | **<0.001** |
| PD-Prophylaxis vs. PD-Untreated | 0.89 [0.78-1.02] | 0.098 |  | 0.81 [0.66-0.98] | **0.030** |
| **>=40% eGFR decline** |  |  |  |  |  |
| PD-Treated vs. PD-Untreated | 0.71 [0.61-0.83] | **<0.001** |  | 0.69 [0.56-0.84] | **<0.001** |
| PD-Prophylaxis vs. PD-Untreated | 0.72 [0.58-0.90] | **0.004** |  | 0.60 [0.43-0.82] | **0.002** |
| **Incident Albuminuria** |  |  |  |  |  |
| PD-Treated vs. PD-Untreated | 0.91 [0.84-0.99] | **0.030** |  | 0.88 [0.79-0.99] | **0.032** |
| PD-Prophylaxis vs. PD-Untreated | 0.80 [0.71-0.91] | **<0.001** |  | 0.79 [0.67-0.94] | **0.008** |
| **Cohort B - Patients Without Periodontal Disease** |  |  |  |  |  |
|  | **Univariable Analysis** |  |  | **Multivariable Analysis\*\*** |  |
| **Incident CKD** |  |  |  |  |  |
| Regular Dental Prophylaxis (>=1 visit/year) | 0.86 [0.80-0.92] | **<0.001** |  | 0.87 [0.78-0.96] | **0.006** |
| **>=40% eGFR decline** |  |  |  |  |  |
| Regular Dental Prophylaxis (>=1 visit/year) | 0.70 [0.63-0.79] | **<0.001** |  | 0.76 [0.65-0.90] | **0.001** |
| **Incident Albuminuria** |  |  |  |  |  |
| Regular Dental Prophylaxis (>=1 visit/year) | 0.84 [0.79-0.89] | **<0.001** |  | 0.85 [0.78-0.93] | **<0.001** |

\* Adjusted for age, gender, baseline eGFR, observation time, race/ethnicity, body mass index, socioeconomic variables, current smoking/alcohol use disorder, charlson comorbidity index, coronary artery disease and proton pump inhibitors.
\*\* Adjusted for age, baseline eGFR, observation time, race/ethnicity, body mass index, socioeconomic variables, angiotensin converting enzyme inhibitors or angiotensin receptor blockers (ACE-I/ARB), statins, smoking/alcohol, charlson comorbidity index and hypertension
**PD-Treated** – Periodontal Disease with Periodontal Treatment
**PD-Prophylaxis** – Periodontal Disease without Periodontal Treatment but with regular dental prophylaxis (>=1 visit/year)
**PD-Untreated** – Periodontal Disease with neither Periodontal Treatment nor regular dental prophylaxis (<1 visit/year)

**Fig 2:**
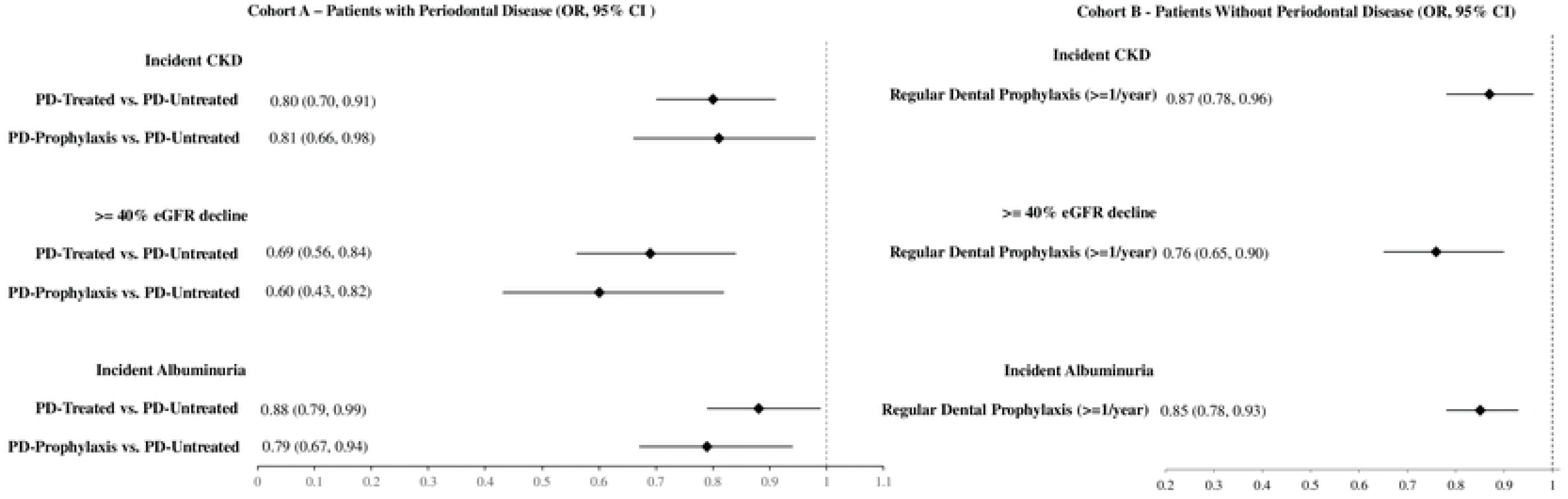
Forest plot of the adjusted logistic regression model for Incident CKD, >=40% eGFR decline and Incident Albuminuria

When further adjusting for baseline UACR (if available), results remained consistent: PD-Treated group (PD-Treated vs. PD-Untreated; OR: 0.75 [0.65-0.87], p<0.001), PD-Prophylaxis vs. PD-Untreated (OR: 0.77 [0.62-0.96], p=0.021).

#### > 40% Reduction in eGFR

Individuals in PD-Treated group or PD-Prophylaxis group had a significantly lower likelihood of experiencing a 40% decline in eGFR compared to PD-Untreated group. Those in PD-Treated group had a 29% lower likelihood of this outcome (95% CI: [0.61-0.83], p<0.001), and those in PD-Prophylaxis group had 28% lower likelihood of this outcome (95% CI: [0.58-0.90], *p*=0.004). (Table 3) After adjusting for other variables, these associations remained consistent (PD-Treated vs. PD-Untreated; OR: 0.69 [0.56-0.84], p<0.001; PD-Prophylaxis vs. PD-Untreated; OR: 0.60 [0.43-0.82], p=0.002). (Fig 2) Other factors associated with this outcome were older age, higher BMI, current or former smoking, higher CCI, Cardiovascular Disease (CVD) including Heart Failure, and statin usage (S3 Table).

When further adjusting for baseline UACR (if available), results remained consistent: PD-Treated group (PD-Treated vs. PD-Untreated; OR: 0.65 [0.52-0.81], p<0.001), PD-Prophylaxis vs. PD-Untreated (OR: 0.62 [0.43-0.88], p=0.009).

### Cohort B

#### Incident CKD

In the non-periodontal disease cohort, those with regular dental prophylaxis (≥ 1 visit/year) had significantly lower odds of incident CKD (Table 2). Regular dental prophylaxis had a 14% lower likelihood of incident CKD (95% CI: [0.80-0.92], *p*<0.001). After adjusting for confounding variables, these associations remained consistent (OR: 0.87 [0.78-0.96], *p*=0.006) (Table 3) (Fig 2). Other variables associated with new-onset CKD were non-white race, ACE/ARB usage, current or former smoking, alcohol use disorder, higher CCI, and hypertension (S4 Table).

When further adjusting for baseline UACR (if available), results remained consistent (OR: 0.86 [0.77-0.97], *p*=0.011).

#### > 40% Reduction in eGFR

Patients with regular dental prophylaxis (≥ 1 visit/year) had a 30% lower likelihood of experiencing a 40% eGFR decline (95% CI: [0.63-0.79], p<0.001). After covariate adjustment, these associations remained consistent (OR: 0.76 [0.65-0.90], *p*=0.001) (Table 3) (Fig 2). Other variables associated with this outcome were older age, ACE/ARB usage, current/former smoking, hypertension, and higher CCI (S4 Table). Analysis of variance inflation factors for all variables included in the models (Cohort A & B) showed that all variables had VIF ≤ 5, indicating no evidence of strong multicollinearity.

When further adjusting for baseline UACR (if available), results remained consistent (OR: 0.85 [0.77-0.93], *p*<0.001).

### Analysis of UACR Labs

Within the entire cohort, 56,206 (65.1%) had evidence of UACR labs. The total number of labs were 831,231 and the average number of UACR labs per-patient was 11 (IQR: 6-16).

### Cohort A

#### Incident Albuminuria

In patients with periodontal disease, those in PD-Treated or in PD-Prophylaxis group had significantly lower odds of developing albuminuria compared to PD-Untreated group. PD-Treated patients had 0.91 times lower odds of albuminuria (95% CI: 0.84–0.99, p=0.030), and PD-Prophylaxis patients had 0.80 times lower odds (95% CI: 0.71-0.91, p<0.001). After adjusting for covariates, these associations remained significant (PD-Treated vs. PD-Untreated; OR (95% CI): 0.88 [0.79-0.99], *p*=0.032; PD-Prophylaxis vs. PD-Untreated; OR (95% CI): 0.79 [0.67-0.94], *p*= 0.008). (Table 3, Fig 2) Other variables associated with albuminuria were older age, non-white race, Hispanic ethnicity, higher BMI, higher CCI, Cardiovascular Disease (CVD) including Heart Failure, and statin usage (S3 Table).

### Cohort B

#### Incident Albuminuria

In the non-periodontal disease cohort, regular dental prophylaxis (≥ 1 visit/year) exhibited significantly lower odds of developing albuminuria.(Table 2) Regular dental prophylaxis was associated with 0.84 times lower odds of albuminuria (95% CI: 0.79 – 0.89, p<0.001). This association persisted after covariate adjustment (adjusted OR: 0.85, 95% CI: 0.78-0.93, p<0.001). (Table 3, Fig 2) Other variables associated with albuminuria were older age, higher BMI, smoking status, ACE-I/ARB usage, hypertension, and higher CCI (S4 Table). Analysis of variance inflation factors for all variables included in the models (Cohort A & B) showed that all variables had VIF ≤ 5, indicating no evidence of strong multicollinearity.

## Discussion

In this large retrospective observational cohort study of patient with normal kidney function at baseline, we showed that untreated periodontal disease is associated with an increased risk of incident CKD, new-onset albuminuria, and rapid eGFR decline over a median follow up of seven years. Additionally, undergoing periodontal treatment and adhering to regular dental visits, including at least one annual dental prophylaxis, is associated with a reduced risk of these outcomes. Even in patients without periodontal disease, regular dental prophylaxis at least once a year is associated with a reduced risk of incident CKD, new-onset albuminuria, and rapid eGFR decline compared to those visiting a dentist less than once a year.

This is the first study that investigated the dental care and kidney outcomes in the large US population with dental benefits, examining patients with and without PD. Findings in our study are particularly meaningful given the robustness of our outcome definition. To define CKD, not only we required to have eGFR < 60 ml/min/1.73m2 but also it must be 25% lower than baseline value and required to be confirmed with additional lab value at least 90 days apart. Furthermore, Rapid eGFR decline of >= 40% is suggestive of rapid CKD progression and widely accepted as a surrogate endpoint for end-stage renal disease (ESRD).[13]

The US Veterans Health Administration (VHA) offers a great opportunity to conduct an in-depth analysis of electronic health records by integrating dental care with medical records. However, eligibility for dental care within the VA is guided by the provisions of regulations Title 38 United States Code (U.S.C.) §§ 1712 and Title 38 Code of Federal Regulations (CFR) §§ 17.160-166. Only veterans categorized as Class I, Class IIA, Class IIC, and Class IV are eligible for continuous and comprehensive dental care at VA. Thus, longitudinal evaluation of these veterans gives us a unique opportunity for outcome analysis without the concern for the affordability of dental care.

Periodontal disease exhibits a complex, bidirectional and pathophysiologically intertwined relationship with CKD. Advanced CKD significantly alters calcium-phosphorus metabolism, compromises immune system, and creates a pro-inflammatory environment that exacerbates oral microbiological dysbiosis. Uremic conditions further exacerbate oral dysbiosis, as salivary urea hydrolyzes to ammonia, elevating pH and fostering a pathogenic biofilm dominated by species such as *Porphyromonas gingivalis*, which promotes gingival inflammation and periodontal pocket formation.[14,15] Conversely, PD serves as a chronic inflammatory focus that contributes to systemic inflammatory cascades, potentially accelerating kidney function deterioration through proinflammatory cytokines and endothelial dysfunction.[9] So, it can be inferred that reducing periodontal inflammation through treatment and maintaining optimal oral health through regular dental prophylaxis may be linked to a reduced inflammatory burden, which in turn could lead to improved kidney outcomes. In our study, by focusing exclusively on patients with normal kidney function, we have eliminated the risk factors associated with CKD that could contribute to the development of periodontal disease.

The observations of this retrospective study suggest that untreated PD may be linked to an increased risk of CKD. The observed improvement in CKD outcomes following therapeutic and prophylactic interventions supports the idea that PD may contribute to CKD. Our analysis of patients without PD shows a strong association between regular dental prophylaxis and better kidney outcomes, suggesting that even the microinflammatory burden from untreated dental plaque could play a role in negative outcomes.

In a small cohort of Prima Indian patients with diabetes, severe periodontal disease (characterized by missing teeth and alveolar bone loss on radiographs) was linked to a two-fold increase in albuminuria incidence and a 3.5-fold increase in ESRD.[6] Similarly, a study of 699 African Americans revealed that severe periodontitis was associated with a fourfold greater risk of developing CKD, but this was not observed with milder forms of periodontal disease.[16] Our study differs from previous research work. Not only do we confirm that untreated periodontal disease is correlated with poorer kidney outcomes in large population with extended follow-up period but we also demonstrate that nonsurgical treatment of periodontal disease is associated with improved kidney outcomes.

Taiwan has implemented a National Health Insurance program that provides free dental prophylaxis every six months. Research of Taiwanese population shows that dental prophylaxis reduces the risk of acute myocardial infarction, stroke, ESRD and mortality.[17–19] Chung et al.’s propensity score matching analysis showed that dental prophylaxis reduces ESRD risk in CKD patients, with a dose-dependent effect.[19] The study evaluated dental scaling within 24 months prior to an ICD-9 CKD diagnosis, comparing patients who received scaling with those who did not. While 60% had PD, non-PD patients showed the greatest benefit from scaling, whereas PD patients showed no significant advantages.

Our study aligns with the findings of Chung et al. However, we focused on longitudinal data from patients with normal kidney function, with a median follow-up of 7 years, during which we collected all reported eGFR (median 14) and UACR (median 11). We employed robust outcome definitions and examined both patients with and without periodontal disease, revealing that periodontal treatment and regular dental prophylaxis are associated with improved kidney outcomes in both groups.

Our study has several limitations. First, due to the study’s retrospective nature, unknown confounding factors may have influenced the results. Second, the diagnosis of periodontal disease is primarily based on ICD codes, however, we used validated codes that were used previously.[20] Third, the lack of data concerning pocket depth, attachment loss, and tooth extractions limits our capacity to accurately assess the severity of periodontal disease. Nevertheless, the ICD-based diagnosis encompasses milder forms of PD; despite this inclusion, we still observed a consistently increased risk of kidney outcomes, underscoring the robustness of our findings. Fourth, our analysis concentrated exclusively on non-surgical periodontal treatment options, specifically scaling and root planing and periodontal maintenance. We did not consider invasive treatments, such as flap surgery, which, while uncommon, may not have significantly influenced the overall outcomes. Fifth, our assessment focused on dental care provided by the VA or its fee basis referral to community providers and did not take into account any private dental care not covered by the VA. Finally, our study cohort consists mainly of male veterans, and further research is needed to make our findings applicable to the general population.

Our study has several strengths. We conducted a longitudinal analysis on a large population eligible for dental care, adjusting for socioeconomic factors, which minimizes the impact of financial constraints on the results. Additionally, we had an extended observation period with multiple laboratory values collected (average 14 eGFR and 11 UACR results). We used robust, well-accepted definitions for kidney outcomes.

## Conclusion

In this large retrospective observational cohort study, we have shown an association between untreated periodontal disease and an increased risk of incident CKD, new-onset albuminuria, and rapid eGFR decline. Our findings underscore the importance of periodontal treatment and regular dental prophylaxis in reducing the risk of these conditions. Even in patients without periodontal disease, regular dental prophylaxis has been shown to be beneficial, reducing the risk of incident CKD, new-onset albuminuria, and rapid eGFR decline compared to individuals who visit a dentist less than once a year.

## Data Availability

Data cannot be shared because it belongs to the U.S. government.

## Acknowledgments

We would like to express our sincere gratitude to the Department of Veterans Affairs, Office of Dentistry, for providing us with eligibility data. However, we want to emphasize that the entire analysis was conducted independently by the authors. The views expressed in this article are solely those of the authors and do not necessarily reflect the official position or policies of the Department of Veterans Affairs or the US government.

This work was supported using resources and facilities of the Department of Veterans Affairs (VA) Informatics and Computing Infrastructure (VINCI), VA HSR RES 13-457.

## Supporting information

**S1 Table: Data Dictionary**

**S2 Table: Cohort A (Periodontal Disease) vs. Cohort B (No-Periodontal disease).**

**S3 Table – Logistic Regression Models, Cohort A (Patients with Periodontal Disease)**

**S4 Table – Logistic Regression Models, Cohort B (Patients without Periodontal Disease)**

## References

1. Eke PI, Thornton-Evans GO, Wei L, Borgnakke WS, Dye BA, Genco RJ. Periodontitis in US Adults: National Health and Nutrition Examination Survey 2009-2014. J Am Dent Assoc. 2018;149: 576–588.e6. doi:10.1016/j.adaj.2018.04.023

2. Scannapieco FA, Gershovich E. The prevention of periodontal disease—An overview. Periodontology 2000. 2020;84: 9–13. doi:10.1111/prd.12330

3. Lockhart PB, Bolger AF, Papapanou PN, Levison ME. Periodontal disease and atherosclerotic vascular disease: does the evidence support an independent association? Circulation. 2012;125: 2520–2544. doi:10.1161/CIR.0b013e31825719f3

4. Veazie S, Vela K, Parr NJ. Evidence Brief: Detection and Treatment of Dental Problems on Chronic Disease Outcomes. Washington, DC 20420: Department of Veterans Affairs, Veterans Health Administration, Health Services Research & Development Service; 2021 Feb pp. 1–59. Available: https://www.hsrd.research.va.gov/publications/esp/TopicNomination.cfm

5. Borgnakke WS, Ylöstalo PV, Taylor GW, Genco RJ. Effect of periodontal disease on diabetes: systematic review of epidemiologic observational evidence. J Periodontol. 2013;84: S135–152. doi:10.1902/jop.2013.1340013

6. Shultis WA, Weil EJ, Looker HC, Curtis JM, Shlossman M, Genco RJ, et al. Effect of periodontitis on overt nephropathy and end-stage renal disease in type 2 diabetes. Diabetes Care. 2007;30: 306–311. doi:10.2337/dc06-1184

7. Chen Y-T, Shih C-J, Ou S-M, Hung S-C, Lin C-H, Tarng D-C, et al. Periodontal Disease and Risks of Kidney Function Decline and Mortality in Older People: A Community-Based Cohort Study. Am J Kidney Dis. 2015;66: 223–230. doi:10.1053/j.ajkd.2015.01.010

8. Valenzuela-Narváez RV, Valenzuela-Narváez DR, Valenzuela-Narváez DAO, Córdova-Noel ME, Mejía-Ruiz CL, Salcedo-Rodríguez MN, et al. Periodontal disease as a predictor of chronic kidney disease (CKD) stage in older adults. J Int Med Res. 2021;49: 3000605211033266. doi:10.1177/03000605211033266

9. Li L, Zhang Y-L, Liu X-Y, Meng X, Zhao R-Q, Ou L-L, et al. Periodontitis Exacerbates and Promotes the Progression of Chronic Kidney Disease Through Oral Flora, Cytokines, and Oxidative Stress. Front Microbiol. 2021;12: 656372. doi:10.3389/fmicb.2021.656372

10. Inker LA, Eneanya ND, Coresh J, Tighiouart H, Wang D, Sang Y, et al. New Creatinine- and Cystatin C–Based Equations to Estimate GFR without Race. New England Journal of Medicine. 2021;385: 1737–1749. doi:10.1056/NEJMoa2102953

11. Sumida K, Nadkarni GN, Grams ME, Sang Y, Ballew SH, Coresh J, et al. Conversion of Urine Protein–Creatinine Ratio or Urine Dipstick Protein to Urine Albumin–Creatinine Ratio for Use in Chronic Kidney Disease Screening and Prognosis. Ann Intern Med. 2020;173: 426–435. doi:10.7326/M20-0529

12. Kim JH. Multicollinearity and misleading statistical results. Korean J Anesthesiol. 2019;72: 558–569. doi:10.4097/kja.19087

13. Levey AS, Inker LA, Matsushita K, Greene T, Willis K, Lewis E, et al. GFR Decline as an End Point for Clinical Trials in CKD: A Scientific Workshop Sponsored by the National Kidney Foundation and the US Food and Drug Administration. American Journal of Kidney Diseases. 2014;64: 821–835. doi:10.1053/j.ajkd.2014.07.030

14. Altamura S, Pietropaoli D, Lombardi F, Del Pinto R, Ferri C. An Overview of Chronic Kidney Disease Pathophysiology: The Impact of Gut Dysbiosis and Oral Disease. Biomedicines. 2023;11: 3033. doi:10.3390/biomedicines11113033

15. Martínez Nieto M, De Leon Rodríguez ML, Anaya Macias R del C, Lomelí Martínez SM. Periodontitis and chronic kidney disease: A bidirectional relationship based on inflammation and oxidative stress. World J Clin Cases. 2024;12: 6775–6781. doi:10.12998/wjcc.v12.i35.6775

16. Grubbs V, Vittinghoff E, Beck JD, Kshirsagar AV, Wang W, Griswold ME, et al. Association Between Periodontal Disease and Kidney Function Decline in African Americans: The Jackson Heart Study. J Periodontol. 2015;86: 1126–1132. doi:10.1902/jop.2015.150195

17. Lee Y-L, Hu H-Y, Chou P, Chu D. Dental prophylaxis decreases the risk of acute myocardial infarction: a nationwide population-based study in Taiwan. Clin Interv Aging. 2015;10: 175–182. doi:10.2147/CIA.S67854

18. Lee Y-L, Hu H-Y, Huang N, Hwang D-K, Chou P, Chu D. Dental Prophylaxis and Periodontal Treatment Are Protective Factors to Ischemic Stroke. Stroke. 2013;44: 1026–1030. doi:10.1161/STROKEAHA.111.000076

19. Chung Y-H, Kuo H-C, Liu H-Y, Wu M-Y, Chang W-J, Chen J-T, et al. Association between Dental Scaling and Reduced Risk of End-Stage Renal Disease: A Nationwide Matched Cohort Study. Int J Environ Res Public Health. 2021;18: 8910. doi:10.3390/ijerph18178910

20. Jurasic MM, Gibson G, Wehler CJ, Orner MB, Jones JA. Caries prevalence and associations with medications and medical comorbidities. Journal of Public Health Dentistry. 2019;79: 34–43. doi:10.1111/jphd.12292

